# Hypokalemia and Clinical Implications in Patients with Coronavirus Disease 2019 (COVID-19)

**DOI:** 10.1101/2020.02.27.20028530

**Authors:** Dong Chen, Xiaokun Li, Qifa Song, Chenchan Hu, Feifei Su, Jianyi Dai, Yinghai Ye, Jianping Huang, Xiaoming Zhang

**Affiliations:** Departments of Infectious Diseases, Wenzhou Central Hospital and Sixth People’s Hospital of Wenzhou, Wenzhou, Zhejiang Province, China; Wenzhou Medical University, Wenzhou, Zhejiang Province, China; Department of Microbiology, Ningbo Municipal Centre for Disease Control and Prevention, Ningbo, Zhejiang Province, China; Department of Microbiology, Wenzhou Municipal Centre for Disease Control and Prevention, Wenzhou, Zhejiang Province, China

**Keywords:** Hypokalemia, severe acute respiratory syndrome coronavirus 2 (SARS-CoV-2), Coronavirus Disease 2019 (COVID-19), Clinical Implication, biomarker

## Abstract

**BACKGROUND:** SARS-CoV-2 has caused a series of COVID-19 globally. SARS-CoV-2 binds angiotensin I converting enzyme 2 (ACE2) of renin–angiotensin system (RAS) and causes prevalent hypokalemia

**METHODS:** The patients with COVID-19 were classified into severe hypokalemia, hypokalemia, and normokalemia group. The study aimed to determine the relationship between hypokalemia and clinical features, the underlying causes and clinical implications of hypokalemia.

**RESULTS:** By Feb 15, 2020, 175 patients with COVID-19 (92 women and 83 men; median age, 46 [IQR, 34–54] years) were admitted to hospital in Wenzhou, China, consisting 39 severe hypokalemia-, 69 hypokalemia-, and 67 normokalemia patients. Gastrointestinal symptoms were not associated with hypokalemia among 108 hypokalemia patients (*P* > 0.05). Body temperature, CK, CK-MB, LDH, and CRP were significantly associated with the severity of hypokalemia (*P*<0.01). 93% of severe and critically ill patients had hypokalemia which was most common among elevated CK, CK-MB, LDH, and CRP. Urine K^+^ loss was the primary cause of hypokalemia. 1 severe hypokalemia patients was given 3 g/day, adding up to an average of 34 (SD=4) g potassium during hospital stay. The exciting finding was that patients responded well to K^+^ supplements when they were inclined to recovery.

**CONCLUSIONS:** Hypokalemia is prevailing in patients with COVID-19. The correction of hypokalemia is challenging because of continuous renal K^+^ loss resulting from the degradation of ACE2. The end of urine K^+^ loss indicates a good prognosis and may be a reliable, in-time, and sensitive biomarker directly reflecting the end of adverse effect on RAS system.

---

Since December 2019, a life-threatening coronavirus was recognized as the etiological factor of a series of severe pneumonia cases arising in the city Wuhan of China ^1^. This virus was named severe acute respiratory syndrome coronavirus 2 (SARS-CoV-2) by the International Committee on Taxonomy of Viruses. The disease was named Coronavirus Disease 2019 (COVID-19) by World Health Organization(WHO). SARS-CoV-2 invades human cells through binding angiotensin I converting enzyme 2 (ACE2) on the cell membrane ^2^. ACE2 is widely distributed in many types of human tissues, especially in the vital organs, such as heart, liver, kidney, and lungs ^3^. ACE2 is viewed as the principal counter-regulatory mechanism for the main axis of renin–angiotensin system (RAS) that is an essential player in the control of blood pressure and electrolyte balance. ^4^. SARS-CoV-2 binds ACE2 and enhances the degradation of ACE2, and thus decreases the counter-act of ACE2 on RAS. The final effect is to increase reabsorption of sodium and water, and thereafter increase blood pressure and excretion of potassium (K^+^) ^5^. Besides, patients with COVID-19 often had gastrointestinal symptoms such as diarrhea and vomiting ^6^. Collectively, the impacts on RAS and gastrointestinal system by COVID-19 probably lead to disruptions of homeostasis of electrolytes and pH. One of the disruptions that may reflect the progression of COVID-19 and should be closely monitored is hypokalemia. K^+^ ions are the predominant intracellular cations and influence membrane polarization of cells. Hypokalemia results in cellular hyperpolarity, increases resting potential, and hastens depolarization in cardiac cells and lung cells ^7^. Severe hypokalemia of under 3 mmol/L plasma K^+^ can trigger ventricular arrhythmia and respiratory muscle dysfunction, both conditions being life-threatening in patients in severe COVID-19 condition. This knowledge implies that hypokalemia may have a consuderable impact on the treatment outcome of patients with COVID-19 and should be seriously tackled as these patients have a high prevalence of dysfunction in heart, lungs, and other vital organs.

As little is known about the prevalence of hypokalemia and its adverse effects on the treatment outcomes of patients with COVID-19, here we reported the high prevalence of hypokalemia in such patients and investigated the relevant causes and clinical meaning. We also aimed to determine the possible relationship between hypokalemia and the treatment outcomes of these patients.

## METHODS

### STUDY POPULATION

This study included patients with COVID-19 who were admitted to the hospital for infectious diseases in the city Wenzhou of China from Jan 11 to Feb 15, 2020. All patients were aged ≥14 years old and were diagnosed to have COVID-19 according to the criteria issued by the National Health and Health Commission of the People’s Republic of China ^8^. All cases were screened with the presence of cough, fever, and radiographic presentations and confirmed by the real-time PCR on the respiratory tract samples or blood samples to test for a sequence of SARS-CoV-2. Based on disease severity, four types of cases were specified according to the criteria mentioned above, i.e., mild cases that showed mild clinical manifestations and no pneumonia; moderate cases that showed respiratory symptoms and mild pneumonia; severe cases that showed pneumonia and any of acute respiratory distress syndrome (ARDS) with respiratory rate of over 30 times/min and oxygen saturation of less than 93%; and critical cases that showed pneumonia and any of shock, respiratory failure, and failures in other organs.

### STUDY DESIGN

This retrospective study was conducted during the COVID-19 outbreak. A trained team of medical staff reviewed and collected the recorded demographic, epidemiological, clinical, and laboratory data in a standardized data collection form modified from electronic medical records. The patients were classified into three groups according to three levels of plasma K^+^, i.e., severe hypokalemia group (under 3 mmol/L), hypokalemia group (3–3.5 mmol/L), and normokalemia group (over 3.5 mmol/L). The clinical features, therapy, and treatment outcomes were compared between three groups, aiming to specify the relationship between hypokalemia and clinical features, and to determine the underlying causes and clinical implications of hypokalemia. To elucidate the mechanisms of hypokalemia, an experimental group of 20 patients with hypokalemia and a control group of 20 patients with normokalemia were established and their K^+^ in point urine sample was measured and compared. In addition, among the patients with COVID-19, plasma K^+^ concentrations were compared between the patients with and without gastrointestinal symptoms. We also randomly selected several clinically mild and severe cases with hypokalemia to trace the treatment effect of K^+^ supplements.

This study was approved by the Ethics Committee of Wenzhou Central Hospital and Sixth People’s Hospital of Wenzhou and followed the Declaration of Helsinki. Written consent was acquired for all patients.

### DATA COLLECTION

The epidemiological investigation was focused on the transmission mode through the history of travel or residence in Wuhan and the close contact with confirmed patients within 14 days according to the WHO interim surveillance recommendations for human infection with novel coronavirus ^9^. The etiological examinations included real-time PCR assays that detected SARS-CoV-2, influenza, respiratory syncytial virus, adenovirus, and para-influenza virus in respiratory specimens. The immunological responses and possible sepsis were evaluated by measuring white blood cells, erythrocyte sedimentation rate, procalcitonin, and C-reactive protein. Lungs, liver, cardiovascular, and renal functions were evaluated using laboratory tests, including coagulation profile, plasma assays (creatinine, blood urea nitrogen, alanine aminotransferase, aspartate transferase, creatine kinase, lactate dehydrogenase, and electrolytes), arterial blood gas examination, and electrocardiogram (ECG). K^+^ in spot urine samples was measured and reported in mmol/gram of creatinine.

Additionally, computed tomography (CT) was used to diagnose infection in the lungs. Therapeutic data and treatment outcomes were also retrieved. The therapeutic principles included general support, monitoring of lungs, liver, kidney and myocardial functions, active control over the high fever, oxygen uptake and K^+^ supplements if necessary, and antiviral therapy with interferon-α and lopinavir/ritonavir. Treatment outcomes were referred to as improved, cured, and failed, and the number of hospital days and the days for patients to have SARS-CoV-2-PCR-negative results. The patients who were discharged from the hospital were required to be quarantined for two weeks.

### STATISTICAL ANALYSIS

Continuous measurements with normal distribution were described in mean and standard deviation (SD), while those that were not normally distributed were described in median and interquartile range (IQR). Categorical variables, such as onsets of abnormal laboratory values, were described in counts of events and percentages. The comparison between groups was conducted using Fisher’s exact test for categorical variables when the numbers of events in both groups were over five, and student test for normally distributed continuous measurements. *P* <0.05 was used as the significance threshold value. All analyses were conducete by SPSS (version 20.0).

## RESULTS

During the study period, 175 patients with COVID-19 were included, consisting of 92 women and 83 men (Table 1). The median age was 46 (IQR, 34–54) years. Fifty-seven (33%) patients had a history of exposure to the epidemic area. The patients consisted of 39 patients with severe hypokalemia (<3 mmol/L), 69 patients with hypokalemia (3–3.5 mmol/L), and 67 patients with normokalemia (>3.5 mmol/L). Only 10 patients had a plasma K^+^ concentration of >4 mmol/L. No statistical difference was identified in demographic features and the prevalence of a history of exposure to the epidemic area between three groups. Seventy-one patients had underlying diseases, including 28 patients with hypertension, 12 patients with diabetes, and 31 patients with other conditions. The prevalence of underlying diseases was associated with the severity of hypokalemia (*P*<0.05).

**Table 1.**
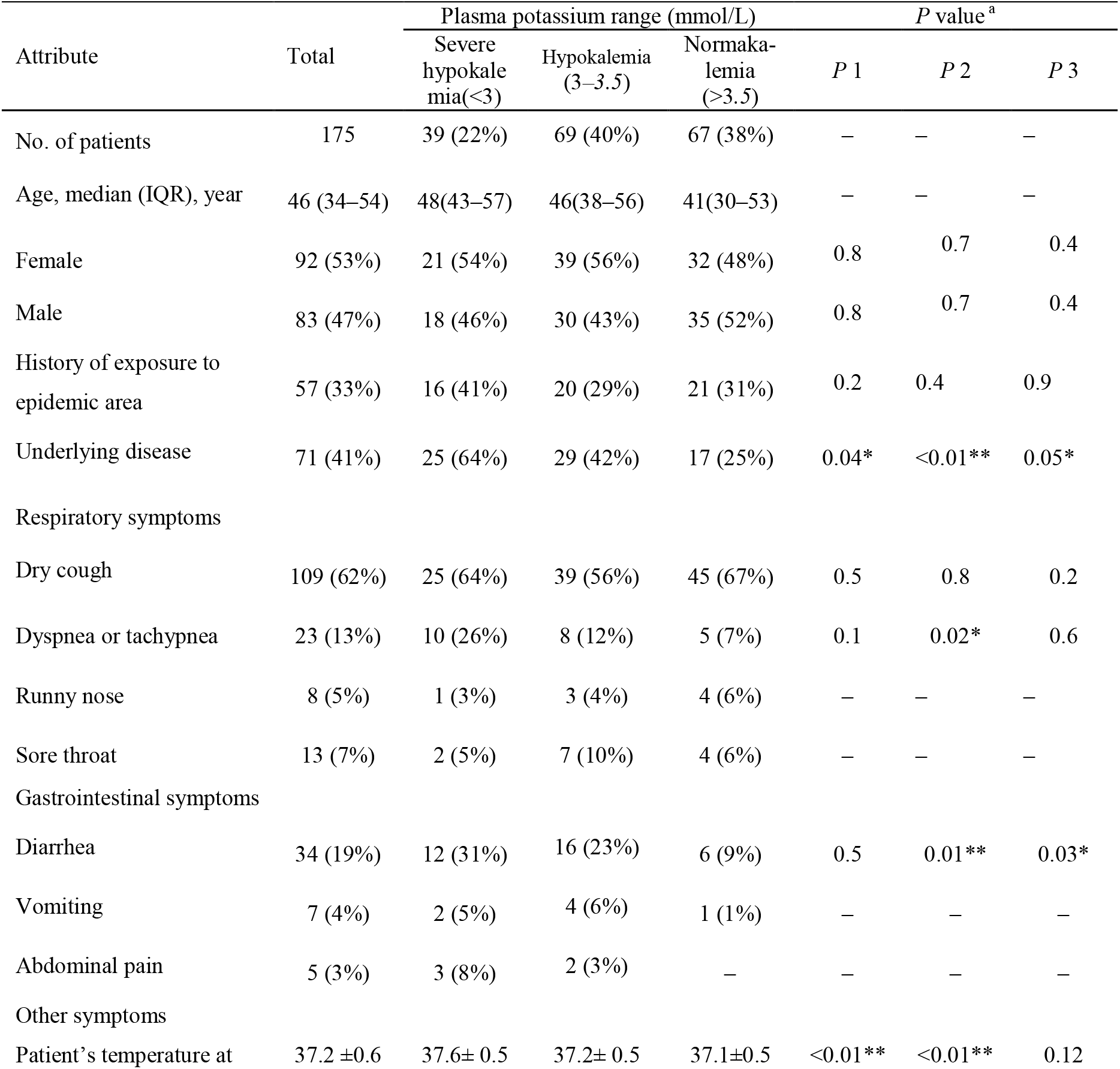
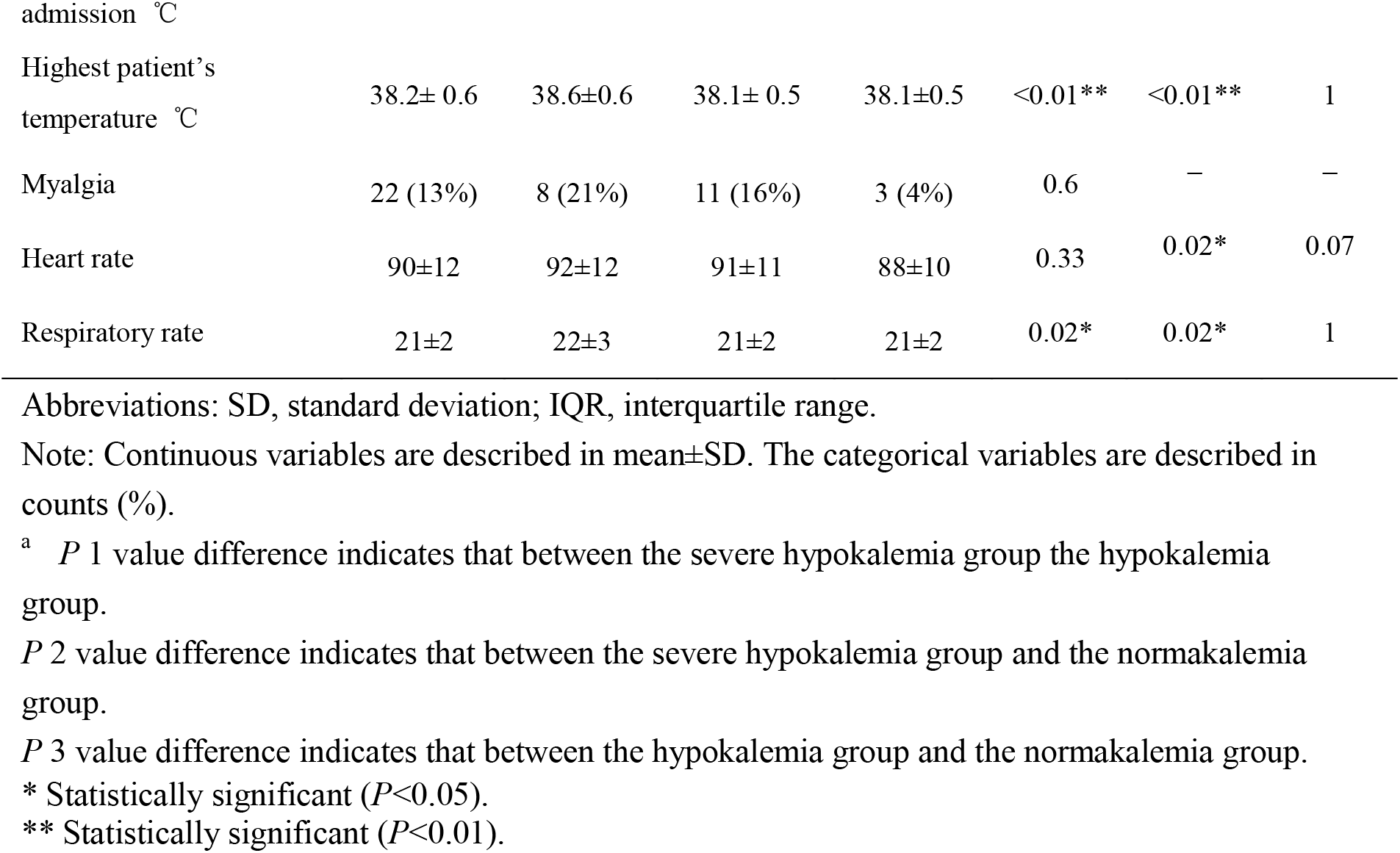
Epidemiological and clinical symptoms of patients stratified by three different levels of plasma potassium

Concerning the presenting symptoms (Table 1), except that the patients with severe hypokalemia had a statistically higher prevalence of dyspnea or tachypnea, no association was observed between the severity of hypokalemia and the prevalence of common respiratory symptoms, such as cough and running nose. As to the gastrointestinal symptoms that might cause hypokalemia, 34 (19%) patients had diarrhea with an average of 6 onset times per day and often ended within 1–4 days, dominantly in the patients with hypokalemia (*P* < 0.05). Few patients had vomiting and abdominal pain. The patients with severe hypokalemia had statistically higher body temperature (mean, 37.6°C; SD, 0.5 °C) than the patients with hypokalemia (mean=37.2°C, SD=0.5 °C, and *P*<0.0) and the patients with normokalemia (mean=37.1°C, SD=0.5°C, and *P*<0.01). Similar difference was also observed in the highest patient’s temperature during the hospital stay. The patients with severe hypokalemia also had a higher heart rate and respiratory rate than the remaining patients (*P* < 0.05).

Among a series of laboratory examinations (Table 2), creatine kinase (CK), CK-MB, lactate dehydrogenase, aspartate transferase, and erythrocyte sedimentation rate were significantly associated with the severity of hypokalemia (*P*<0.01). Collectively, these attributes with elevated values were generally related to myocardial injury. A total of 108 patients with hypokalemia exhibited moderate decreases in sodium, white blood cells, and lymphocyte than the 67 patients with normokalemia. Among these 108 patients, 39 patients exhibiting gastrointestinal symptoms had a mean plasma K^+^ of 3.22 (SD, 0.22) mmol/L, which was not statistically different from the mean value of 3.26 (SD, 0.28) mmol/L in 69 patients without gastrointestinal symptoms. Regarding the urine K^+^ output, 20 patients with hypokalemia had a mean K^+^ concentration of 32 (SD, 11) mmol/gram of creatinine, which was statistically higher than the corresponding mean of 18 (SD, 7) mmol/gram of creatinine for 20 patients with normokalemia (*P<*0.01).

**Table 2.** Laboratory examinations of patients stratified by three levels of plasma potassium

| Attribute | Total | Plasma potassium range (mmol/L) |  |  | *P* value<sup>a</sup> |  |  |
| --- | --- | --- | --- | --- | --- | --- | --- |
|  |  | *Severe hypokalemia* (<3) | *Hypokalemia* (3–3.5) | *Normokalemia* (>3.5) | *P* 1 | *P* 2 | *P* 3 |
| No. of patients | 175 | 39 (22%) | 69 (40%) | 67 (38%) | – | – | – |
| WBC ($\times 10^9$/L) | 4.9±1.5 | 4.6±1.4 | 4.9±1.6 | 5.2±2.1 | 0.16 | 0.05* | 0.17 |
| No. of leukopenia (<4$\times 10^9$/L) | 44 (25%) | 11 (28%) | 16 (23%) | 17(25%) | 0.6 | 0.8 | 0.8 |
| Lymphocyte ($\times 10^9$/L) | 1.28±0.6 | 1.12±0.5 | 1.24±0.6 | 1.39±0.6 | 0.14 | 0.01** | 0.07 |
| No. of hypolymphocytemia (<1.1$\times 10^9$/L) | 61 (35%) | 16 (41%) | 27 (39%) | 18 (27%) | 1 | 0.1 | 0.1 |
| CRP (0–8 mg/L) | 19±21 | 28±23 | 19±20 | 13±18 | 0.4 | 0.1 | 0.06 |
| ESR (0–20 mm/h) | 26±16 | 34±15 | 26±16 | 22±15 | <0.01** | <0.01** | 0.07 |
| CK (U/L) | 96±188 | 131±188 | 95±84 | 78±51 | 0.02* | 0.08 | 0.08 |
| No. of elevated CK (>170 U/L) | 24 (14%) | 15 (38%) | 5 (7%) | 4 (6%) | <0.01** | <0.01** | 1 |
| CK-MB (U/L) | 20±13 | 35±18 | 21±14 | 17±12 | <0.01** | <0.01** | 0.04* |
| No. of elevated CK-MB (>18 U/L) | 39 (22%) | 25 (64%) | 7 (10%) | 7 (10%) | <0.01** | <0.01** | 1 |
| LDH (U/L) | 216±66 | 256±81 | 213±60 | 196±60 | <0.01** | <0.01** | 0.05* |
| No. of elevated LDH (>240 U/L) | 57 (33%) | 23 (59%) | 18 (26%) | 16(24%) | <0.01** | <0.01** | 0.8 |
| ALT (<40 U/L) | 31±28 | 37±35 | 30±30 | 28±30 | 0.14 | 0.08 | 0.35 |
| AST (<40U/L) | 31±18 | 41±20 | 30±19 | 26±14 | <0.01** | <0.01** | 0.08 |
| Creatinine (25–70 umol/L) | 68±31 | 71±17 | 69±31 | 65±33 | 0.35 | 0.14 | 0.27 |
| BUN (3–7 mmol/L) | 3.7±1.2 | 3.9±1.3 | 3.7±1.3 | 3.6±1.4 | 0.22 | 0.14 | 0.33 |
| pH | 7.42±0.04 | 7.43±0.04 | 7.42±0.03 | 7.41±0.03 | 0.07 | <0.01** | 0.03 |
| No. of pH>7.45 | 19 (11%) | 11 (28%) | 5 (7%) | 3 (4%) | <0.01** | – | – |
| pCO<sub>2</sub> | 5.4±0.8 | 5.5±0.7 | 5.4±0.9 | 5.3±0.8 | 0.27 | 0.10 | 0.24 |
| Oxygen saturation (93–100%) | 96.6±2.2 | 96.3±2.4 | 96.6±2.0 | 96.8±2.1 | 0.24 | 0.13 | 0.28 |
| Potassium (3.5–5.5 mmol/L) | 3.4±0.4 | 2.9±0.2 | 3.4±0.2 | 3.8±0.3 | <0.01** | <0.01** | <0.01** |
| Sodium (135–145 mmol/L) | 138±3 | 136±3 | 138±3 | 138±3 | <0.01** | <0.01** | 1 |
| Computed tomography (CT) and electrocardiogram ( ECG) examinations |  |  |  |  |  |  |  |
| Pulmonary ground-glass opacities | 174(100%) | 39(100%) | 68(100%) | 67(100%) | 1 | 1 | 1 |
| ECG | 63 (36%) | 30 (77%) | 23 (33%) | 10 (15%) | <0.01** | <0.01** | <0.02* |

Abbreviations: CK, creatine kinase; LDH, lactate dehydrogenase; ESR, erythrocyte sedimentation rate; CRP, C-reactive protein; ALT, alanine aminotransferase; AST, aspartate transferase; BUN,
blood urea nitrogen.
Note: See Table 1.

In the part of CT and ECG examinations, nearly all patients had pulmonary ground-glass opacities, in trast, abnormal ECG results were significantly correlated with the severity of hypokalemia (*P*<0.01) as the prevalence of abnormal ECG results was 77%, 33%, and 15% for patients with severe hypokalemia, hypokalemia, and normokalemia, respectively.

In terms of severity, there were 37 severe cases and 3 critically cases (Table 1 and Table 4). Severe and critically ill patients had higher occurrence of severe hypokalemia, hypokalemia, abnormal ECG presentations, as well as elevated CK, CK-MB, LDH, and CRP (all *P*<0.01). Among these abnormal indices, hypokalemia was most common as 93% of severe and critically ill patients had hypokalemia. In contrast, elevated BUN and creatinine were rare. Abnormal ECG presentations were usually representative of hypokalemia and improved with K^+^ supplement treatment. Elevated CK, CK-MB, and LDH often came to normal level within 3–6 days (Figure 1). Elevated ALT and AST were generally mild and came to normal level after liver support therapy (Table 2 and Table 4). Most patients with intermittent abnormal oxygen saturation were improved after oxygen inhalation except for three critically ill patients who were given invasive mechanical ventilation.

**Table 3.** Therapy and outcomes of patients stratified by three levels of plasma potassium

| Attribute | Total | Plasma potassium range (mmol/L) |  |  | *P* value<sup>a</sup> |  |  |
| --- | --- | --- | --- | --- | --- | --- | --- |
|  |  | *Severe hypokalemia* (<3) | *Hypokalemia* (3–3.5) | *Normokalemia* (>3.5) | *P* 1 | *P* 2 | *P* 3 |
| No. of patients | 175 | 39 (22%) | 69 (40%) | 67 (38%) | – | – | – |
| Oxygen inhalation | 51 (29%) | 16 (41%) | 24 (35%) | 11 (16%) | 0.5 | 0.01** | 0.02* |
| Interferon-α | 170 (97%) | 39(100%) | 67 (97%) | 64 (96%) | 1 | 1 | 1 |
| Lopinavir/ritonavir | 151 (86%) | 39(100%) | 62 (90%) | 50 (75%) | 0.05* | <0.01** | <0.01** |
| Arbidol | 151 (86%) | 36 (92%) | 62 (90%) | 53 (79%) | 1 | 0.10 | 0.10 |
| Hormone | 12 (7%) | 7 (18%) | 5 (7%) | – | 0.1 | – | – |
| Complication | 11 (6%) | 6 (15%) | 3 (4%) | 2(3%) | – | – | – |
| Severe cases | 37 (21%) | 17 (44%) | 14 (20%) | 6 (9%) | 0.01* | <0.01** | 0.2 |
| Critically ill cases | 3 (2%) | 3 (8%) | – | – | – | – | – |
| No. of discharged patients | 52 (30%) | 12 (31%) | 22 (32%) | 18 (27%) | 1 | 0.7 | 0.6 |
| Days of hospital stay<sup>b</sup> | 16±6 | 19±8 | 16±6 | 15±5 | 0.01 | <0.01** | 0.14 |
| Days to turn PCR-negative<sup>b</sup> | 13±7 | 14±6 | 13±7 | 11±5 | 0.22 | <0.01** | 0.03* |

Note: See Table 1.
<sup>b</sup>The values are calculated according to the number of the patients who were cured and discharged.

**Table 4.** Occurrence of abnormal biomarkers between the group of severe and critically ill cases and the group of moderate and mild cases

| Attribute | Total | Severe and critically ill | Moderate and mild | *P* value |
| --- | --- | --- | --- | --- |
| No. of patients | 175 | 40 | 135 | – |
| K<sup>+</sup> (mean±SD) | 3.4±0.4 | 3.1±0.3 | 3.5±0.4 | <0.01** |
| K<sup>+</sup> (<3 mmol/L) | 19 (11%) | 15 (38%) | 4 (3%) | <0.01** |
| K<sup>+</sup> (<3.5 mmol/L) | 78 (44%) | 22 (55%) | 56 (41%) | 0.1 |
| CK (>170 U/L) | 24 (14%) | 11 (28%) | 13 (10%) | <0.01** |
| CK-MB (>18 U/L) | 39 (22%) | 24 (60%) | 15 (11%) | <0.01** |
| LDH (>240U/L) | 57 (33%) | 26 (65%) | 31 (23%) | <0.01** |
| BUN (>7 mmol/L) | 1 (1%) | 1 (2%) | – | – |
| Creatinine (>110 umol/L) | 1 (1%) | 1 (2%) | – | – |
| Oxygen saturation (<93%) | 8 (5%) | 8 (20%) | – | – |
| CRP (0–8 mg/L) | 85 (49%) | 32 (80%) | 53 (39%) | <0.01** |
| ALT (<40 U/L) | 34 (19%) | 13 (33%) | 21 (16%) | 0.02* |
| AST (<40U/L) | 28 (16%) | 12 (30%) | 16 (12%) | 0.01** |
| No. of leukopenia (<4×10<sup>9</sup>/L) | 44 (25%) | 14 (35%) | 30 (22%) | 0.1 |
| ECG | 63 (36%) | 25 (62%) | 38 (28%) | <0.01** |

Note: See Table 1.

**Figure 1.**
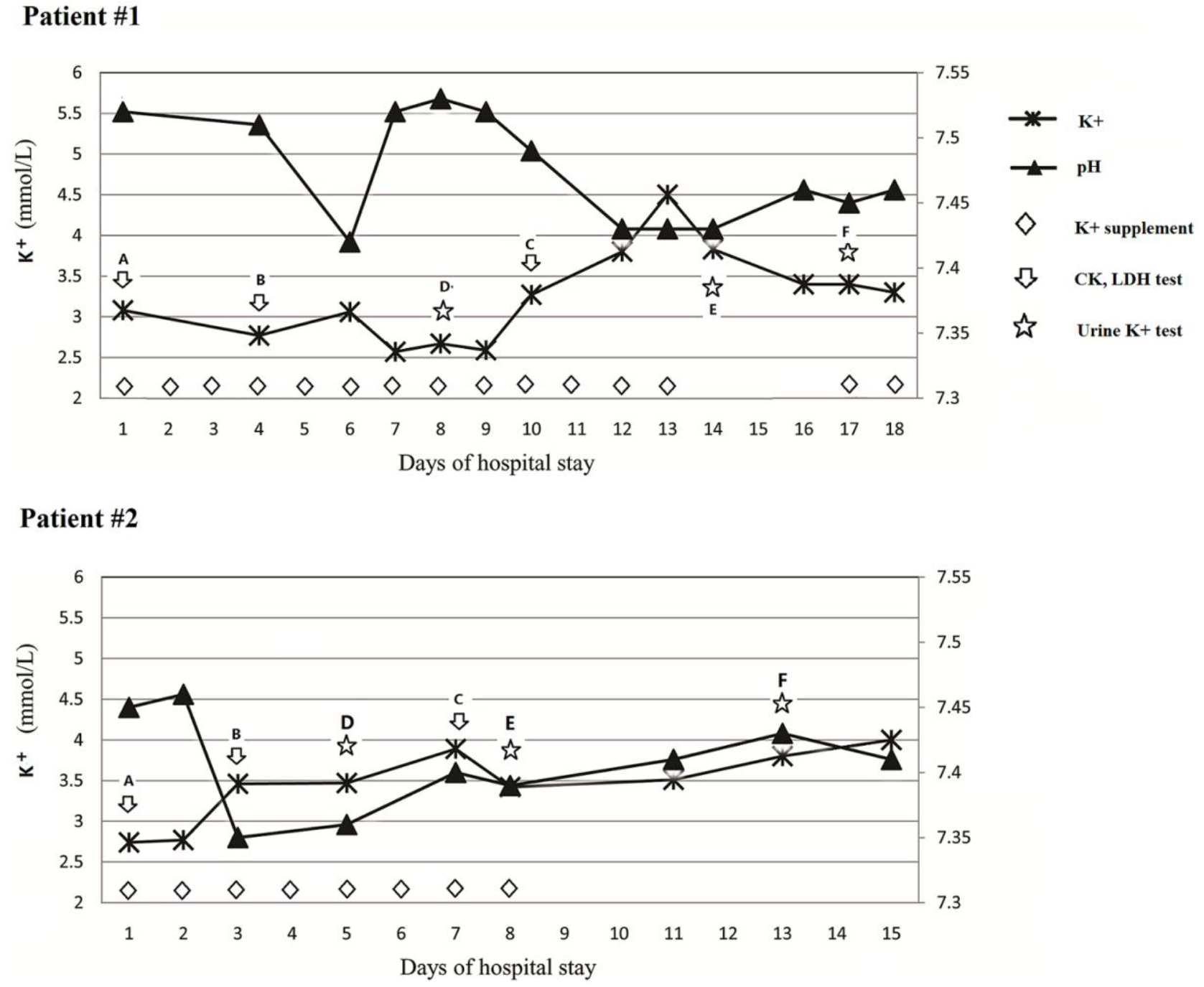
Trend in plasma K^+^ and pH in COVID-19 progression and the patient’s response to K^+^ supplement. Patient #1: The figure shows CK and LDH fall into the normal range in a few days. The relapse of elevated loss of urinary K^+^ leads to the failure in K^+^ supplements. Time point A–C: CK, 530, 365, 40 U/L; CK-MB, 65, 45, 11 U/L; LDH, 357, 270, 212 U/L, respectively. Time point D–F: Urine K+, 41, 50, 38 umol/gram of creatinine, respectively. Patient #2: The figure shows CK and LDH fall into the normal range in a few days. The loss of urine K^+^ is alleviated, which ensures the effective treatment of K^+^ supplements. Time point A–C: CK, 258, 132, 62 U/L; CK-MB, 35, 26, 12 U/L; LDH, 355, 238, 220 U/L, respectively. Time point D–F: Urine K+, 32, 21, 9 umol/gram of creatinine, respectively.

Turning to the therapy and treatment outcomes (Table 3), the severity of hypokalemia meant a higher prevalence of severe cases (*P*<0.01) and necessitated more frequent usage of antiviral drugs such as lopinavir/ritonavir (*P*<0.01) and arbidol, as well as oxygen inhalation (*P*<0.05) besides interferon-α. Till the time when the manuscript was written, 62 (35%) patients were cured and discharged from the hospital. The severity of hypokalemia led to substantially more days of hospital stay (*P*<0.05) and more days needed for patients to have PCR-negative results (*P*<0.05). As to the K^+^ supplements treatment, the average amount of K^+^ supplements for 21 discharged patients with severe hypokalemia was 3 g/day, adding up to an average of 34 (SD, 4) g during the whole hospital stay for each patient.

To be noteworthy, the most significant adverse effect of hypokalemia is a myocardial injury that can be well reflected by ECG, CK, and CK-MB. A careful check identified a noticeable jump of increase in the prevalence of abnormal ECG results and elevated CK and particularly elevated CK-MB in the patients with severe hypokalemia (Table 2).

## DISCUSSION

The current study illustrated the high prevalence of hypokalemia in the patients with COVID-19 and the positive association between the degree of hypokalemia and the severity of COVID-19. The study also proved that hypokalemia was more attributable to renal loss of K^+^ than gastrointestinal loss.

Previous literature has proved that sufficient and appropriate levels of plasma K^+^ have a protective role in preventing myocardial failure through weakening cellular hyperpolarity and depolarization ^10,11^. SARS-CoV-2 can result in heart dysfunction due to the intensively expressed ACE2 in the patients’ myocardial cells that act as receptors for this virus. Therefore, it is beneficial to patients that plasma K^+^ levels be frequently checked and maintained between 4.0 and 5.5 mmol/L or 4.5 and 5.5 mmol/L for serum K^+^ as serum generally has more K+ than plasma ^12^. In the present study, hypokalemia was prevailing among the patients with COVID-19, up to 62% (108/175) patients having plasma K^+^ that was under 3 mmol/L. Among 175 patients, only 10 patients had plasma K^+^ of 4 mmol/L recommended for patients with myocardial dysfunction. That strikingly low prevalence of optimal concentration of plasma K^+^ implied a massive risk for the patients’ heart to be affected by both hypokalemia and the virus. As an emerging infectious disease, the available data on serum K^+^ levels is scanty. Only one previous study on 41 patients reported 4.2 mmol/L as the mean serum K^+^, as well as higher K^+^ in the ICU patients (mean, 4.6 mmol/L) than non-ICU patients (mean, 4.1 mmol/L), suggesting that the elevated serum K^+^ was associated with severity of illness ^13^. However, our findings that were derived from more patients were contrary to this previous study. Considering the implications of serum K^+^ concentrations in this disease, further investigation is necessary.

The present study also proved that the degree of hypokalemia was correlated with several clinical features reflecting the severity of the disease. These features included the underlying conditions, high body temperature, and notably, the elevated laboratory indices related to myocardial injury, such as CK, CK-MB, lactate dehydrogenase, aspartate transferase, and erythrocyte sedimentation rate, as well as abnormal ECG results. The other indices, including WBC, lymphocyte, CRP, ALT, were correlated with hypokalemia with less strength than the attributes mentioned above. Regarding the blood gas results, the higher prevalence (28%) of pH of over 7.45 was seen in the patients with severe hypokalemia because severe hypokalemia led to alkalosis due to H^+^-K^+^ exchange between intracellular and extracellular fluid ^14^. Nevertheless, the prevalence of abnormal oxygen saturation and CO_2_ pressure were not sensitive enough to identify the difference between patients with different K^+^ levels. The patients showing renal dysfunction were rare according to a generally normal concentration of blood urea nitrogen and creatinine, as well as sufficient urine output. The globally normal renal function ensured the safe application of K^+^ supplements. Notably, because of the variety in different laboratories and assays that may produce various reference values, it is not recommended to compare results directly. The current study also listed the number and percentage of several abnormal laboratory results, such as WBC, CK, CK-MB, etc, which were considered to be important indices for hypokalemia, myocardial injury, and evaluation of disease. We expected these arrangements of results might facilitate future studies.

Seeing that hypokalemia was prevalent among patients with COVID-19 and associated with the severity of the disease, elucidation of the mechanisms for hypokalemia was elementary for understanding and treatment of COVID-19. In the current situation, two probable causes of hypokalemia are increased gastrointestinal loss and increased urinary loss ^15^. Both causes can find their way in the patients with COVID-19, as mentioned in the introduction section. However, the present findings indicated that gastrointestinal loss might not contribute much to hypokalemia as based on the following reasons: only a small proportion of patients with hypokalemia showing gastrointestinal symptoms, e.g., 31% of patients with severe hypokalemia having diarrhea; among the patients with hypokalemia, no significant difference between those with and those without diarrhea; the average number of diarrhea onsets being 5 times/day and diarrhea ending in a short time, meaning that the diarrhea was mild. Therefore, hypokalemia might principally result from increased urine loss. This was proved in this study by the dramatically increased urine K^+^ output in the hypokalemia group than the control group with normokalemia. This finding that increased urine K^+^ was the primary cause of hypokalemia was consistent with the pathogenesis of SARS-CoV-2. This virus drives RAS towards enhanced ACE–Ang II (angiotensin II)–Ang II type 1 receptor (AT1) axis by degrading ACE2 that is the principal counter-regulatory mechanism for the central axis of the RAS^4^. The final effect of this disturbance of RAS is the increased distal delivery of sodium and water to collecting tubule of the kidney and enhanced potassium secretion. This effect is similar to the effect of aldosterone that stimulates water and sodium reabsorption and potassium excretion and thus increases body water and blood pressure ^16^.

As hypokalemia has an adverse effect on myocardial functions, in-time treatment is required for achieving good outcomes. However, the mechanism of hypokalemia in the present study hinted that it was challenging to achieve normokalemia in the presence of continuous renal loss of K^+^. This worry was proved by the elevated amount of urine K^+^ measured in umol/gram of creatinine (Figure 1). The continual K^+^ supplements had very little effect on the return of normokalemia when the urine loss of K^+^ persisted in the severe cases. Interestingly, we found a noteworthy phenomenon in most patients who were cured of COVID-19. As shown in Figure 1, when patients were inclined to recovery, they responded well to K^+^ supplement treatment and steadily returned to normokalemia. This phenomenon indicated the end of urine loss of K^+^ due to disordered RAS balance; in other words, the ACE2 function began to return. This exciting finding suggested that return of normokalemia might be a reliable biomarker for monitoring ACE2 function.

Although COVID-19 causes injury to lungs, heart, liver, and kidney, our study showed the occurrence of abnormal indices related to heart, liver, and kidney was low, and oxygen saturation often returned to normal level upon oxygen inhalation in most patients (Table 2 and Table 4). Several laboratory indices, such as elevated CK, CK-MB, LDH, ALT and AST were usually came to normal level or substantially improved after relevant treatment. The superficial mildness contradicted with the sudden progression of disease in some patients. This contradiction might result from the fact that the biomarkers were not sensitive to reflect the whole progression of this disease. Therefore, a more sensitive biomarker to monitor the ongoing condition is urgently needed for these patients. As discussed above, 93% severe and critically ill patients had hypokalemia, showing that depletion of K^+^ was prevailing. Based on analysis of the trend in plasma K^+^ and urine output of K^+^, the end of the depletion often suggested a good prognosis (Figure 1). Therefore, comprehensive analysis of K^+^ depletion can be achieved by monitoring urine K^+^ loss, plasma K^+^, and the response to K^+^ supplement treatment. Importantly, the information from this analysis related to hypokalemia directly reflects the very basis of the pathogenesis of SARS-CoV-2 and might be a reliable, int-time and sensitive biomarker to reflect the progression of COVID-19.

The present study had some limitations. Most patients were still hospitalized. Only a small proportion of cases were used to evaluate the treatment outcomes. However, from several patients who had been cured, the principal results related to hypokalemia treatment were consistent, which could provide reliable information. Since this life-threatening disease is still ongoing in China and is developing in several countries, we expect our findings can provide timely information about better understanding and treatment of COVID-19.

To summarize, the present study has identified the prevailing hypokalemia in patients with COVID-19. The correction of hypokalemia is challenging because of continuous renal loss of potassium resulting from the degradation of ACE2 by the binding of SAR-CoV-2. The end of loss of K^+^ often indicates a good prognosis and may be a reliable, in-time, and sensitive biomarker that reflects the end of adverse effect on the RAS system by SAR-CoV-2. Due to the possible injury to cardiovascular functions, neurohormonal activation, and other vital organs by hypokalemia, clinicians should pay great attention to hypokalemia and the patients’ response to K^+^ supplements.

## Data Availability

Yes. All data are included in the manuscript.

## ACKNOWLEDGMENTS

This study was supported by Key scientific and technological innovation projects of Wenzhou (ZY202004) and Natural Science Foundation of Ningbo (2017A610273).

All authors declare no interest.

